# Gender Relations, Women Empowerment and Maternal Health Care in sub-Saharan Africa: A Bayesian Multilevel Analysis

**DOI:** 10.1101/2022.09.10.22279809

**Authors:** Simona Simona

## Abstract

Maternal health care utilization still remains crucial in ensuring good pregnancy outcomes and a reduction in maternal and child mortality especially in developing countries. Although several studies have been conducted to investigate determinants of maternal health care utilization, the innfluence of gender relations and women empowerment on maternal healthcare in cross-national context has received little attention. This paper sought out to examine the influence of gender relations and women empowerment on maternal healthcare utilization in sub-Saharan Africa. The analysis pools sample data of 245,955 respondents from the most recent Demographic and Health Surveys (DHS) and World Development Indicators of 35 sub-Saharan African countries. Separate Bayesian multilevel logistic regression models are fitted for adequate antenatal care and institutional delivery in relation to several factors indicating gender relations and women empowerment at three levels. Several components of gender relations and women empowerment were significantly associated with maternal health care after adjusting for covariates. In addition, significant between country and between community variations in the odds of maternal health care utilisation were observed. These results underscore the importance of prioritising contextual gender equity and women empowerment to achieve better utilisation of maternal healthcare services and subsequently, a reduction in maternal mortality in sub-Saharan Africa.

## Background

About 302,000 women die due to maternal-related causes in developing countries, accounting for 99% of the global maternal deaths estimates. An estimated 546 (66%) Maternal Mortality Rates (MMR) are registered within sub-Saharan Africa (sSA) (Alkema et al., 2016). Lack of adequate maternal health care services such as antenatal care and skilled delivery are some the most important factors exacerbate maternal and child mortality (Filippi et al., 2006; Onah et al., 2006; Zureick-Brown et al., 2013). Access to maternal healthcare services helps provide health information that is necessary for healthy pregnancy outcomes (Birmeta et al., 2013) and ensures timely management and treatment of pregnancy complications to minimize maternal deaths (Tey and Lai, 2013). Despite the importance of maternal healthcare in ensuring the safety of both the mother and child, many women in disadvantaged populations still face challenges using these key services. In low-income countries for example, an estimated 52% of women benefit from skilled care during childbirth compared to high-income countries where almost all women have adequate antenatal care and are attended to by skilled health professionals (Alkema et al., 2016).

Factors accounting for inadequate utilization of maternal healthcare services are complex and range from micro to macro-level structures within society. Underlying social, cultural and economic conditions in which a women and her family live are significant determinants of maternal healthcare (Cresswell et al., 2020; Lange et al., 2019; Simona, 2022; Simona, 2020). Among these conditions, gender relations and women empowerment have emerged to be important themes in maternal, sexual and reproductive health discourses (Adjiwanou and LeGrand, 2014; Kane et al., 2016).Systems of social stratification within the social structure tend to disproportionately privilege men with power, status and better access to resources compared to women (Springer et al., 2012). Women experience their reproductive and maternal healthcare within these frames of gendered social relations, which have implication on the nature and quality of care they receive.

Gender relations are also shaped by agency, which reflects a person’s freedom to pursue and achieve goals important to them (Kabeer, 1999; Sen, 1985). Women’s agency is negotiated within societal power structures in specific spheres of life. Agency leads to women empowerment when it’s exercise undermines and challenges power structures which perpetuate the subordination of women (Hanmer and Klugman, 2016; Kabeer, 1999). Lack of empowerment leads to a sustained denial of equality and self-determination. Women empowerment is often related to reproductive rights, making decisions in the family, labour force participation, economic opportunities, basic education and access to health care (Alkire, 2008; Sen, 1999). The assumption is that the presence of these factors will undermine negative structural influence on women and thus improve chances of adequate maternal health care and subsequently reduce maternal mortality.

### Gender relations and maternal health care

Gender is basically relational in nature because men and women gain their social identities and power in relation to one another (Calasanti, 2010). Gender relations are dynamic constructed power relations between men and women institutionalised by social processes to which people orient themselves to gender ideals (Calasanti, 2010; West and Fenstermaker, 1995). Almost all societies are arranged on the basis of gender such that popular ideals of manhood and womanhood result from and affirm gender division of labour. Socially constructed gender division of labour as well as intrinsic social norms define role expectations, obligations and relationships between men and women (Nankinga et al., 2016). These social formulations and norms are reinforced by sanctions embedded within social systems to ensure conformity and dissuade deviance (Blanc, 2001). Examples of inequalities in gender relations considered in previous research include gender division of labour, gender norms, polygamous family types, intimate partner violence, sexual empowerment and practices that limit women’s movements and interactions with other people [Adjiwanou and LeGrand (2014); (Singh et al., 2015, 2012); Nankinga et al. (2016); Kritz and Makinwa-Adebusoye (1999)]. Most of these studies in sub-Saharan Africa trace gender inequalities in the patriarchal superstructure that has the aim of (re)producing male dominance and the subordination of women (Adjiwanou and LeGrand, 2014).

Inequalities in gender relations are often detrimental to access and utilization of maternal healthcare both directly and indirectly. Gender division of labour, which is so defined by society, assigns women with exclusive responsibility to nurture maternal roles. In most cases this means being solely responsible for pregnancy and childbirth-related issues. In situations where women occupy subordinate positions and have limited access to economic resources, this responsibility is consequential to maternal healthcare outcomes. This is an indirect pathway between societal gender norms and use of maternal health care (Yamin et al., 2015). A much more direct association is that of gender ideologies limiting access to economic, social and cultural capital to enable them to properly use the maternal health care continuum, from antenatal care through skilled delivery care to postnatal check-ups for mothers and newborn babies (Adjiwanou and LeGrand, 2014). Gender relations reflect women’s position relative to that of men in society especially in terms of resource control, sexuality and reproduction. As explained by Adjiwanou and LeGrand (2014), the control of women and girls’ sexuality and reproduction is at the core of unequal gender relations and is central to the denial of equality and self-determination of women.

Evidence shows that gender relations both between men and women are strongly associated with health outcomes (Bottorff et al., 2011). Researching the influence of gender relations on any health or health care outcomes is a recognition of the importance of gender dynamics and circumstances under which they interact to influence opportunities and constraints. It is for this reason that there is need for increased attention to gender relations in health research (Bottorff et al., 2011), particularly, unequal gender relations in terms of individual and structural factors that limit human agency. Inequalities in gender relations as permissive gender norms regarding domestic violence, family types and household headship are important variables in this regard.

Norms regarding condoning violence perpetrated by intimate partners have a direct effect on maternal health care utilization by limiting mobility, control over resources and inducing perceptions of vulnerability and loss of self-control in victims (Adjiwanou and LeGrand, 2014; Sarkar, 2008). Family type is a function of culture and it represents polygyny or the practice of men marrying more than one wife. This practice has been associated with poor health outcomes for women and children (Amey, 2002; Bove and Valeggia, 2009). This is because polygyny exacerbates vulnerability to sexually transmitted diseases and mental health problems due to permission of a multiplication of sexual partners and emotional neglect respectively (Bove and Valeggia, 2009). A few studies on household headship on the other hand, indicate that women from female-headed households are more likely to have better health outcomes compared to those from male-headed households (Adhikari and Podhisita, 2010). This is plausible because household heads have control over resources and if it is women, it is expected that they would channel resources towards better health care.

### Women empowerment and maternal healthcare utilisation

There are many variants of women’s empowerment definitions but the most commonly used are those of (Sen, 1995) and (Kabeer, 1999) which both define the term as the acquisition of capabilities to make decision by those who were previously denied those capabilities. This definition reflects the presence of constraining power that has to be overcome in order for the capabilities to materialize. Decision-making autonomy about one’s life and control over resources are key to empowerment. There are different other terms that have been used to refer to empowerment including agency, women’s status and autonomy (Pratley, 2016). Agency refers to the individual’s ability to exercise power or freedom of choice to pursue and achieve that which they value, free from external constraints (Bhattacharya, 2006; Sen, 1985). Women’s status is measured relative to others and it can be defined as respect given to women and powers available to them (Bloom et al., 2001). Autonomy on the other hand, is mostly synonymous with agency, it is the ability to make independent decisions, to manipulate and control the environment (Bloom et al., 2001). Some scholars also see autonomy as the ability to obtain information and using it to make decisions that affect the outcomes of their families and society (Dyson and Moore, 1983; Hanmer and Klugman, 2016).

The social structure with its norms and values governing social behaviour, sometimes allow for certain outcomes to be obtained and for others to be reproduced regardless of agency (Kabeer, 1999). When this happens, freedom of choice is constrained, and certain individuals may obtain or experience the outcome which they do not value or desire. Women who fail to realize their goals have deep rooted structural constraints on their ability to make choices and this reflects lack of empowerment, agency or autonomy. For example, women may desire to deliver in an institution but if they do not have resources to use for transportation to a health facility, then they lack empowerment. Empowered women have access to resources to enable them to achieve their desired goals.

There are several dimensions of empowerment and it is more meaningful to discuss them in relation to Bourdieu’s capital theory, that is, social, economic and cultural dimensions. In Bourdieu’s theory, social dimensions would mean access to social resources through membership to certain networks, reliance on social support and close relations with friends and family members (Malhotra et al., 2005; Pratley, 2016). Economic dimension regards control over material in form of money or property that can easily be leveraged for health purposes (Kabeer, 1999; Malhotra et al., 2005). Cultural dimension of empowerment has to do with access to symbolic and information resources to be used as the basis for action and this may include but limited to educational attainment(Dyson and Moore, 1983).

Some of the dimensions of empowerment have been used to examine the influence of various structural, social, economic and demographic factors on maternal health care health care in sub-Saharan Africa (Simona et al., 2022). This study extends existing literature by distinguishing the conceptualisation of women empowerment from gender relations and examining the relationship between both concepts and maternal health care utilisation. Additionally, the study used three-level Bayesian multilevel models to assess the extent of across country and across community variations in the odds of maternal health care utilisation in sub-Saharan countries. In so doing, the paper examined the relative importance of contextual level factors (community and country-level) and individual level factors in determining maternal health care utilisation. This is important for bolstering context-based interventions aimed at enhancing maternal health care.

## Methods

### The Data

The individual and community-level analysis (level 1 and 2) pools data from 35 Demographic and Health Surveys (DHS) conducted between 2006 and 2015 in sub-Saharan Africa. Countries included are Angola, Benin, Burkina Faso, Burundi, Cameroon, Chad, Congo, Congo DR, Cote d’Ivoire, Ethiopia, Gabon, Gambia, Ghana, Guinea, Kenya, Lesotho, Liberia, Madagascar, Malawi, Mali, Mozambique, Namibia, Niger, Nigeria, Rwanda, Sao Tome and Principe, Senegal, Sierra Leone, Swaziland, Tanzania, Togo, Uganda, Zambia and Zimbabwe. The inclusion criterion for countries was availability of comparable DHS data on maternal healthcare variables (antenatal care visits, delivery care and postnatal care for mothers and newborn babies). The country-level (level 3) data are drawn from World Development Indicators (WDI) of the World Bank Databank. The sample was restricted to selected DHS countries.

The Demographic and Health Surveys (DHS) are nationally representative population-based cross sectional surveys for men and women designed to provide information on several measures including fertility, family planning, mortality, nutrition, maternal and child health, HIV/AIDS, domestic violence and other health indicators. This study utilizes data for 245,955 women in the reproductive age group of 15-49 years. The DHS sample is typically representative at national level, for urban and rural areas, the regional level and sometimes at state/provincial or district level. The surveys have large sample sizes (usually between 5,000 and 30,000 households) and are typically conducted about every 5 years. The DHS respondents are selected using probabilistic two-stage cluster sampling techniques using the most recent population census as a sampling frame in each participating country (Fabic et al., 2012; Fisher and Way, 1988).

World Development Indicators (WDI) are the primary World Bank collection of development indicators, compiled from officially-recognized international sources. These are the most current and accurate global development data available and they provide national, regional and global estimates (World Bank, 2018). For the purposes of this study, data series from 35 out of 48 sub-Saharan African countries, are selected corresponding to countries with available DHS data. Three Country-level variables were selected from the World Bank Databank including the countries’ human development index (HDI), gender inequality index and national female literacy rate.

### Outcome variables

Two separate dichotomous outcome variables of antenatal care (ANC) visits and institutional delivery care were used to measure maternal healthcare utilization. Both outcome variables were derived from the DHS data. Antenatal care in the DHS was measured by the question which asks women how many times they received antenatal care during pregnancy for their most recent birth. The variable was recoded into a binary, based on the World Health Organisation (WHO) recommendations of at least four ANC visits. Thus, no to three antenatal care visits was recoded as “0”, and at least four antenatal care visits was recoded as “1”. For delivery care, women were asked about their place of delivery for the most recent birth. The variable was recoded to either home “0” or institutional delivery “1”. Only the most recent birth was considered and all previous births were excluded. The preference for the most recent birth was because information on maternal health care tends to be more accurate for most recent births compared to that given for other previous births (Kistiana, 2009)

### Individual-level explanatory factors

Gender relations in this study are represented by several intersecting variables reflecting gendered inequalities, ideologies and practices and these include violence partner acceptance (under such reasons as burning food, refusing sex, visitation without permission, arguing and neglecting children), family type (existence of co-wives), female-headed households (yes or no) and gender inequality which is measured at the country level. On the other hand, variables that were used as proxies of women empowerment included household wealth (recoded as first, second, middle, fourth and highest), educational attainment status (no education, primary, secondary or higher), sexual autonomy (ability to refuse sex demands from a partner), employment status (unemployed, self-employed and formally employed), insurance coverage (yes or no), media exposure (having access to a radio, newspapers or television), decision-making autonomy (composite variable measured by the ability to make independent decisions or together with partner on visiting family members, own health care, household purchases and household earnings) and country-level female literacy rate. Maternal age, distance to health facility and human development index are control factors.

### Community-level explanatory factors

The DHS does not capture variables that can describe the characteristics of the communities. The primary sampling units (PSUs) or clusters were used to assess the community or neighborhood context in this study. Community factors have been calculated by aggregating individual level variables within their clusters. PSUs are used to represent communities and neighborhoods because they are the most consistent measure of communities across all DHS surveys and many previous studies have defined communities in a similar manner (Adjiwanou et al., 2018; Ononokpono et al., 2013; Wiysonge et al., 2012; Yebyo et al., 2015). Just like at the individual level, community level factors were defined to reflect the gender relations and the extent of woman’s empowerment within communities. The aggregates were computed using the mean values of the proportions of women in each category of a given individual variable. The aggregate values of clusters were categorised into groups of ‘Lower’ and ‘Higher’ proportions based on national median values. Community education has three categories of ‘lower’, ‘middle’ and ‘higher’ while place of residence retains the original categorization of ‘rural’ and ‘urban’.

### Country-level explanatory factors

The country level variables also indicate gender relations and empowerment empowerment represented by the country’s gender inequality index and female literacy rate respectively. Gender inequality index is an ordinal variable measured on a scale of 1 to 6, 1 being the most equal country and 6 being the worst. Female literacy rate is the percentage of female literacy levels relative to the female population aged 15 and above in a country. The human development index is the composite indicator of life expectancy, education and per capital income. Countries are ranked on a scale of 0 to 1, 0 being the lowest HDI and 1 being the highest. All country-level measurements were for the year 2015 and standardised to the mean of 0 and standard deviation of ‘1’ for uniformity and easy interpretation.

### Statistical analysis

The distribution of respondents by independent and outcome variables was assessed and expressed in percentage form before the analysis was done. In view of the hierarchical structure of the DHS data, whereby individuals are nested within clusters which are in turn nested within countries and my aim to examine the relative variance of use of maternal healthcare at different levels, I used multilevel modelling techniques for the main analysis. Multilevel techniques are suited for this purpose because, in addition to capacity to assess the fixed associations between variables, they recognize the existence of data hierarchies by allowing for residual components at each level in the hierarchy. In this regard, the residual variance is divided into the variance of the group-level residuals and that of the individual-level residuals. The group residuals represent unobserved group characteristics that affect individual-level outcomes. It is these unobserved variables which lead to correlation between outcomes for individuals from the same group (Bryk and Raudenbush, 1992; Goldstein, 2011; Kreft and De Leeuw, 1998; Snijders and Bosker, 2011).

Multilevel logistic regression models are used in this study to examine the probability *p*_*ijk*_ of a woman *i* in the community *j* and country *k* having adequate maternal healthcare utilization. This analysis is represented as:

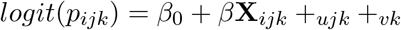

where **X**_*ijk*_ is the vector of explanatory variables at individual, community and country levels, _*ujk*_ is normally distributed with variance 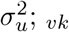 is normally distributed with 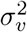

In terms of variances used to understand the relative importance of the general contextual factors of community and country-level characteristics, I used the median odds ratios (MOR) and the variance partition coefficients (VPC). The MOR is on the same scale as the odds ratios and is interpreted as the median value of the odds ratios between individuals from units at high or low risk when randomly choosing 2 individuals from different units. In this study, that would be the odds of having inadequate utilization of maternal healthcare that are determined by unexplained factors at the community and country levels.

The VPC provides information on the share of the variance at each level of operation. The VPC at each level was calculated using the latent method. It assumes a threshold model and approximating the level-1 (individual) variance by *π*^2^*/*3 (≈ 3.29) (Dundas et al., 2014; Goldstein, 2011; Merlo et al., 2005; Rodriguez and Goldman, 2001).

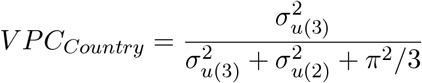

and

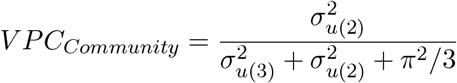

A three-level multilevel model for each of the two outcome variables (ANC visits and delivery care was specified with the three-level structure. Estimates for parameters were obtained using the Bayesian Markov chain Monte Carlo (McMC) Methods in MLwiN through the R2MLwiN package (Zhang et al., 2016) in R. MLwiN uses a combination of Gibbs sampling and Metropolis-Hastings sampling to extract samples from the posterior distribution of unknown parameters. McMC methods allow for specification of the prior distribution which is then combined with the likelihood function produced by the data to create the posterior distribution. McMC methods do not aim to find simple point estimates for the parameters of interest as in frequentest likelihood methods. Instead, they make a large number of simulated random draws from the joint posterior distribution of all parameters and use the draws to make a summary of the underlying distributions (Browne, 2015; Gill, 2014). From these random draws, it is then possible to calculate the posterior mean and standard deviation (SD), as well as density plots of the complete posterior distribution and quantiles of this distribution allowing for the construction of credible intervals. The Bayesian inference with McMC methods are preferred for this analysis because they produce unbiased estimates according to Browne (2015).

Because of limited background information and lack of related previous studies, this study uses non-informative uniform prior distributions with large variances (mean = 0, variance = 106) for regression parameters and inverse gamma (0.001, 0.001) for precision parameters. One chain was specified running for 55,000 iterations with a burn-in length of 5,000 iterations in order to achieve convergence. The convergence of chains was assessed by inspection of trace and auto-correlation plots as shown in the appendix. The Bayesian Deviance Information Criterion (DIC) was used to evaluate the goodness of fit of the models (Browne, 2015; Gelman and Hill, 2007; Lynch, 2007).

### Ethical consideration

This study is based on existing DHS data and publicly available world development indicators. Both have fewer ethical implications because they do not include any identifier information. The DHS surveys are approved by the Institutional Review Board of the ICF International in Calverton, Maryland, USA and specific ethics committees in participating countries. The surveys are conducted by well-trained research assistants who administer informed consent before respondents are interviewed and information is obtained anonymously and confidentially. The data is stored by the DHS Program based in Maryland United States and are publicly available for research purposes upon approval from the program.

## Results

### Sample characteristics

Table 1 presents sampled countries, year of completion of data collection, final sample per country, number of communities in a country, median number of respondents per community and range of respondents in a community. As indicated above, the surveys were conducted between 2006 and 2015. The total number of respondents per country ranged between 1,445 Sao Tome and Principe and 20,192 Nigeria. The number of communities in the sample ranged from 104 for Sao Tome and Principe and 1612 for Kenya. The median number of respondents per community is between 7 and 21. Table 2 presents descriptive statistics for the pooled sample data of 245,955 respondents in SSA at individual level.

**Table 1:** Description of DHS data

| Country | Year | N | Communities | Median N | Range |
| --- | --- | --- | --- | --- | --- |
| Angola | 2015-16 | 8947 | 625 | 15 | 1-26 |
| Benin | 2011-12 | 9111 | 750 | 12 | 2-31 |
| Burkina Faso | 2010 | 3960 | 210 | 17 | 2-49 |
| Burundi | 2010 | 4916 | 376 | 13 | 4-21 |
| Cameroon | 2011 | 7655 | 580 | 13 | 4-21 |
| Chad | 2014-15 | 11104 | 626 | 18 | 3-40 |
| Comoros | 2012 | 2016 | 252 | 08 | 1-16 |
| Congo | 2011-12 | 6463 | 384 | 17 | 4-39 |
| Congo DR | 2013-14 | 11293 | 540 | 21 | 9-38 |
| Cote d'Ivoire | 2011-12 | 5431 | 352 | 14 | 4-37 |
| Ethiopia | 2011 | 7764 | 650 | 13 | 1-26 |
| Gabon | 2012 | 4143 | 336 | 12 | 1-34 |
| Gambia | 2013 | 5385 | 281 | 17 | 2-72 |
| Ghana | 2014 | 4294 | 427 | 09 | 1-33 |
| Guinea | 2012 | 4999 | 300 | 16 | 6-41 |
| Kenya | 2014 | 14949 | 1612 | 09 | 1-25 |
| Lesotho | 2014 | 2596 | 400 | 06 | 1-17 |
| Liberia | 2013 | 5348 | 322 | 16 | 5-32 |
| Madagascar | 2008-09 | 8569 | 600 | 14 | 4-31 |
| Namibia | 2013 | 3974 | 600 | 07 | 1-18 |
| Niger | 2012 | 7680 | 480 | 16 | 3-39 |
| Nigeria | 2013 | 20192 | 904 | 20 | 3-55 |
| Rwanda | 2014-15 | 5955 | 492 | 12 | 3-23 |
| Sao Tome | 2008-09 | 1445 | 104 | 12 | 3-48 |
| Senegal | 2010-11 | 8151 | 392 | 20 | 5-47 |
| Sierra Leone | 2013 | 8524 | 435 | 19 | 5-43 |
| Swaziland | 2006-07 | 2136 | 275 | 07 | 1-18 |
| Tanzania | 2015-16 | 7050 | 608 | 11 | 1-18 |
| Togo | 2013-14 | 5016 | 330 | 14 | 2-34 |
| Uganda | 2011 | 4909 | 712 | 12 | 1-25 |
| Zambia | 2013-14 | 9353 | 722 | 12 | 1-26 |
| Zimbabwe | 2015 | 4833 | 400 | 12 | 1-25 |

**Table 2:**
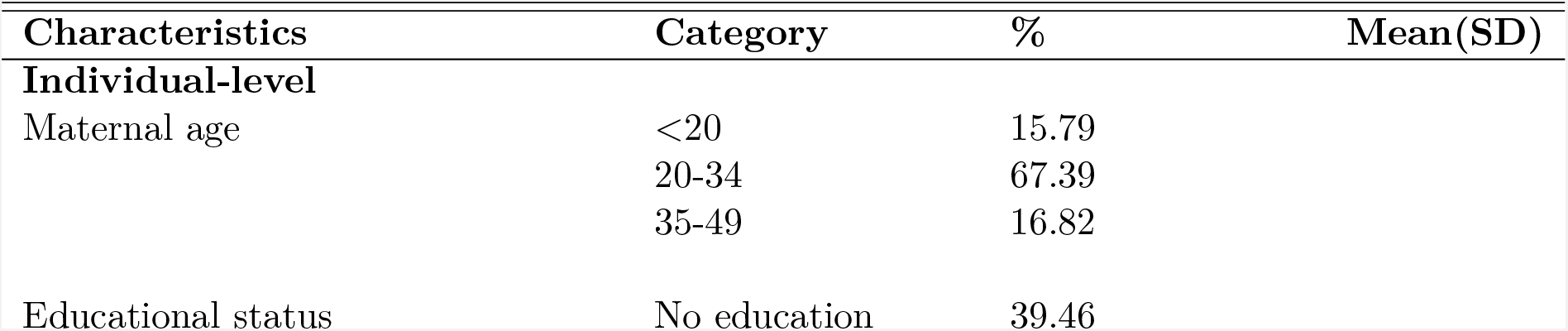
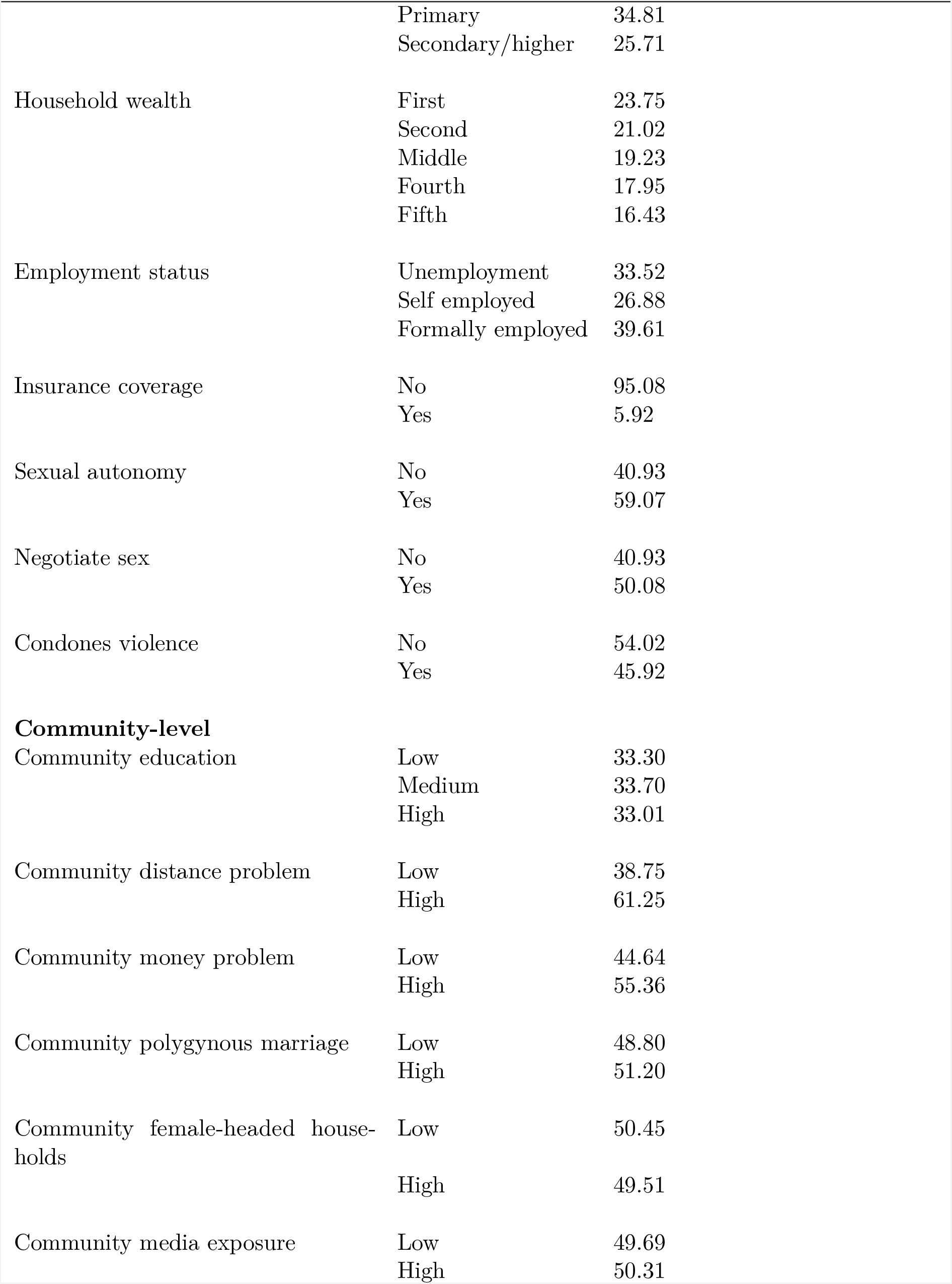
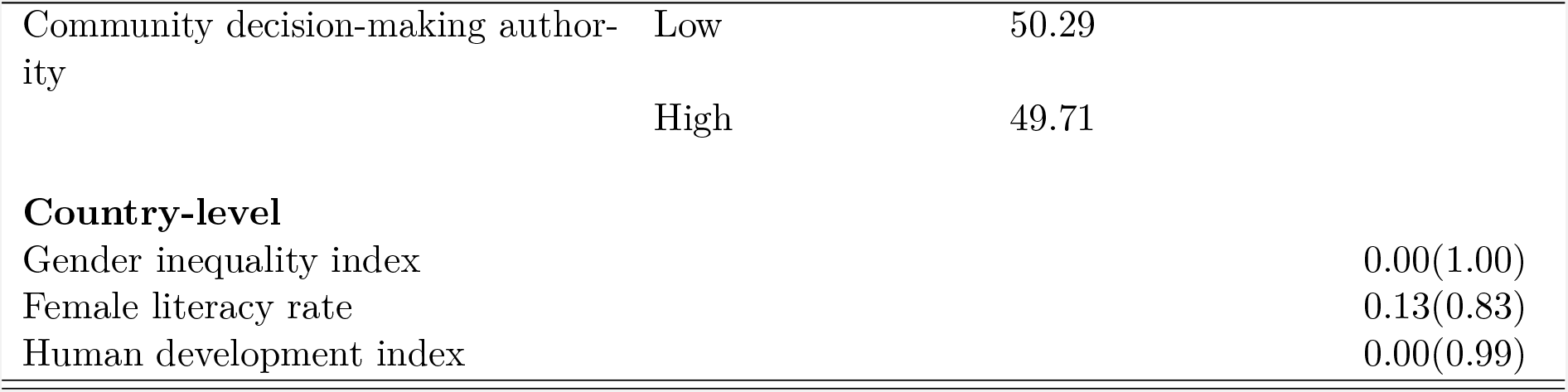
Descriptive statistics for analysis pooled sample (N = 245, 955)

Table 2 shows summary statistics of the explanatory variables included in the analysis. The results show that most of the respondents were aged between 20 and 34 (67.39%), had no education (39.46%) and were in the poorest quintile of the population (23.75). One-third of women were unemployed while 39.61% were in formal employment with 26.88% being employed in the agricultural sector. As expected, the larger majority had no health insurance coverage (95.08%). In terms of sexual autonomy, most of the respondents (59.07%), reported that they were able to refuse sex while a slight majority (50.08) were comfortable to ask their partners to use a condom during a sexual encounter. Slightly over half reported that the use of violence against women is justifiable under certain circumstances.

The community level factors indicate that the proportion of women living in rural areas in sSA is more (68%) than those residing in urban areas. Community education seems to be evenly distributed as the communities with low, medium and higher education are all approximately 33%. The table also indicates that the proportion of women in communities saying that the distance to health facilities is a problem is more (61.25%) than those don’t find it problematic. Similarly, the proportion of those who find money to be a problem in access to health facilities are more (55.36%) than those who don’t. The proportion of women living in communities with high polygamous marriages is more than (51.38%) those living in communities without polygamous marriages and the proportion of women living in communities with low female-headed households is almost equal to that of women living in communities with high female-headed household holds. In both media exposure and decision-making authority, there is almost an equal proportion of women residing in both categories of each variable.

Table 2 also shows the summary statistics of factors measured at the country level in terms of standardised means and standard deviations. Standardization of continuous variables is important to aid the interpretation of relationships in the regression analysis. The results show that the gender inequality index and human development index have means approaching and standard deviations closer to 1, resembling that of standardised normal distribution. Country female employment rate has a standardised mean of 0.13 and standard deviation of 0.83.

Figure 1 reports the distribution of the two outcome variables by country. The results indicate the prevalence of institutional delivery in sSA range between approximately 16% to about 84% while that of ANC visits is between 22% and 86%. The variability in the outcome factors is about the same.

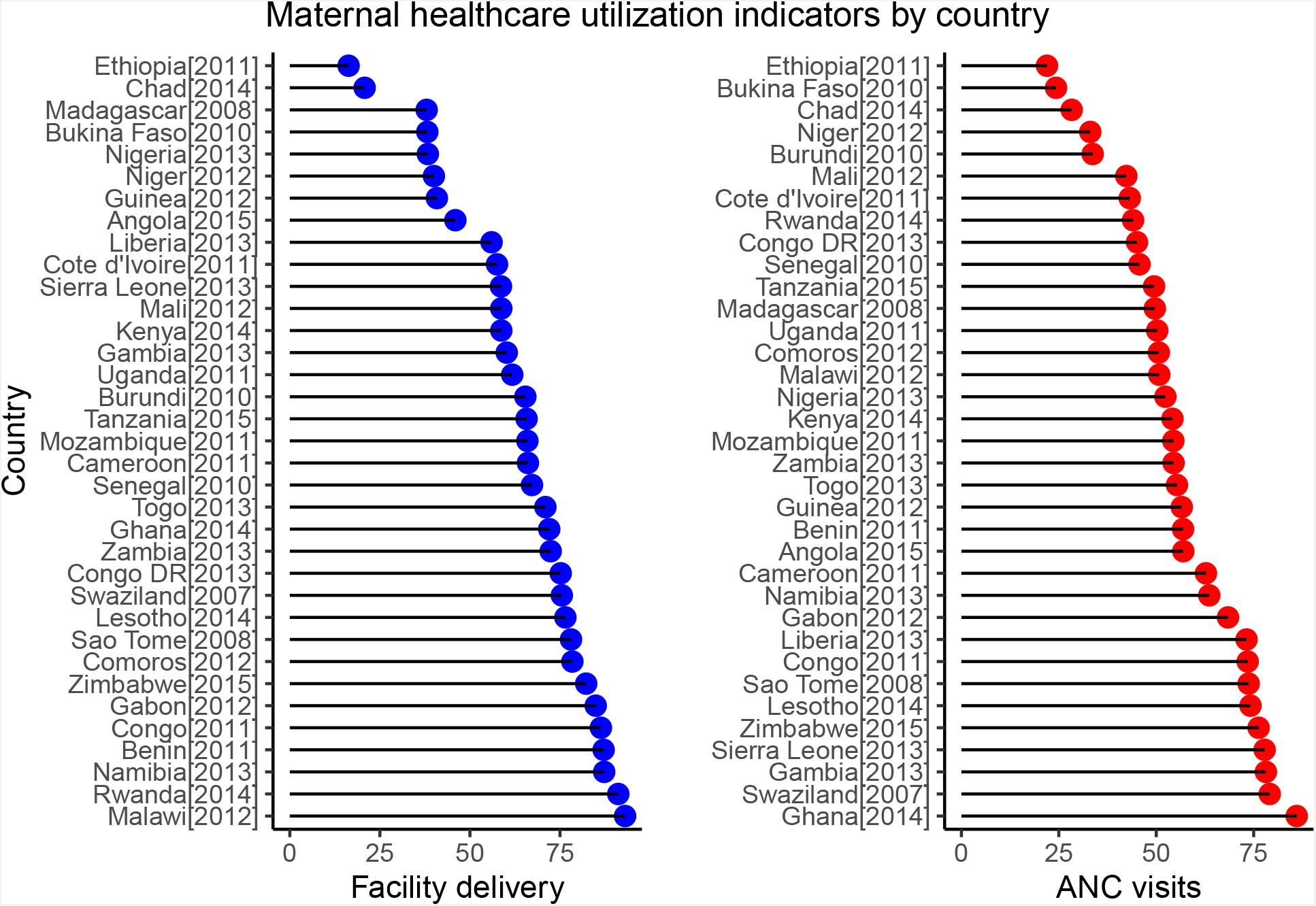

The figure shows that the performance of sSA countries in the use maternal healthcare depends on the specific indicator of maternal healthcare. For antenatal care, Ghana is the best performing country where more than 80% of pregnant women attended antenatal care in their most recent pregnancy. Ghana is followed by Swaziland, Sierra Leone and the Gambia averaging between 77% and 80%. For institutional delivery, we see that Malawi, Rwanda and Namibia are the best performing countries. In terms of least performing countries, Ethiopia, Burkina Faso and Chad seem to be consistently recording low level of maternal healthcare utilization across the continuum of antenatal care and institutional delivery.

### Multilevel analysis

Four models were specified for each of the outcome variables and are reported in table 3 and 4. Model 1 is the null or empty model which contains only the intercept and outcome variable. Model 2 includes the individual level variables (maternal age, educational status, household wealth, employment, sexual autonomy, insurance coverage and condoning violence). Model 3 includes community-level variables (community education, community decision-making authority, community distance to health facility problem, community polygamous marriages, community female-headed households, community money problem and area of residence) and Model 4 has the country-level variables (gender inequality index, national female literacy rate and human development index).

**Table 3:**
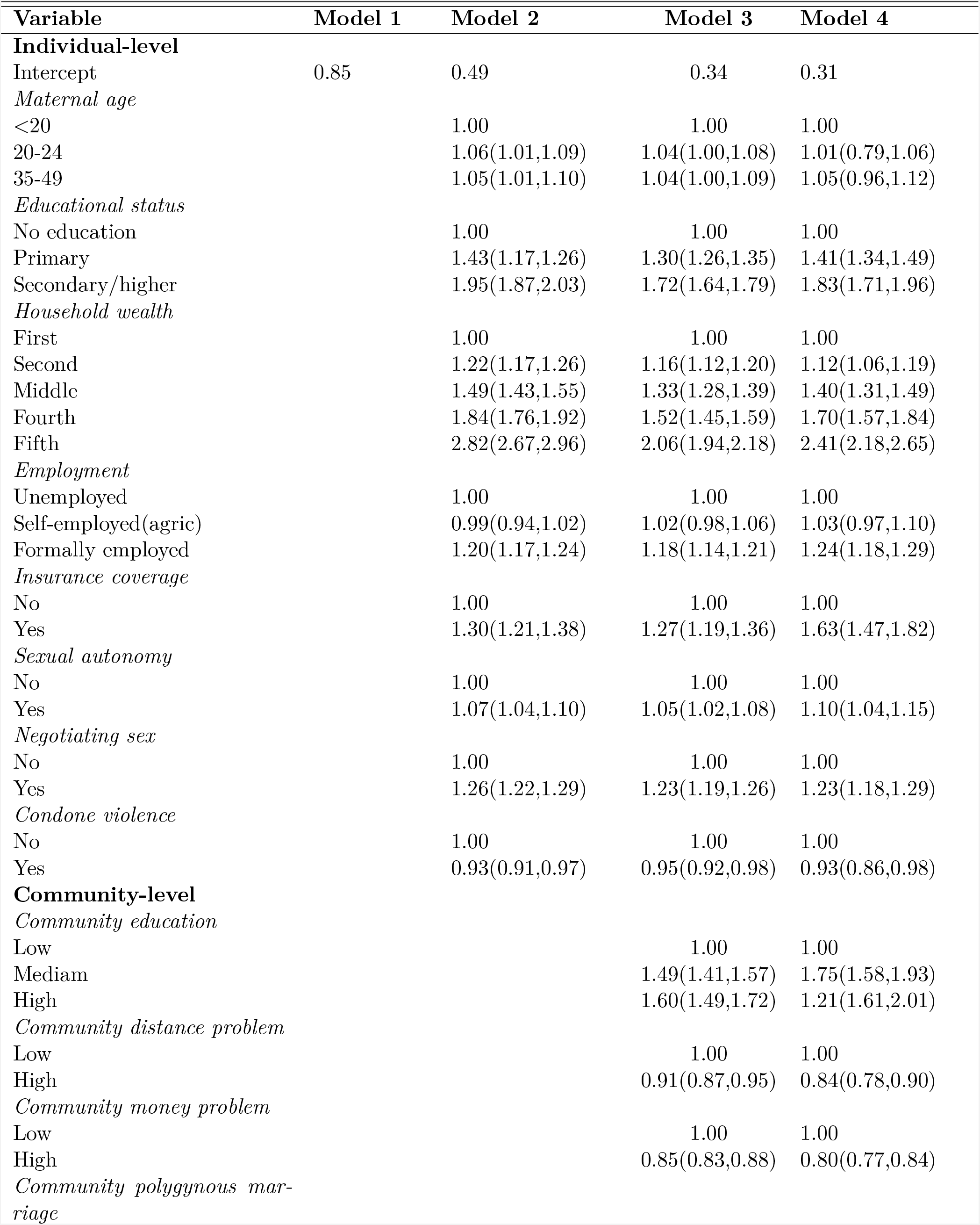
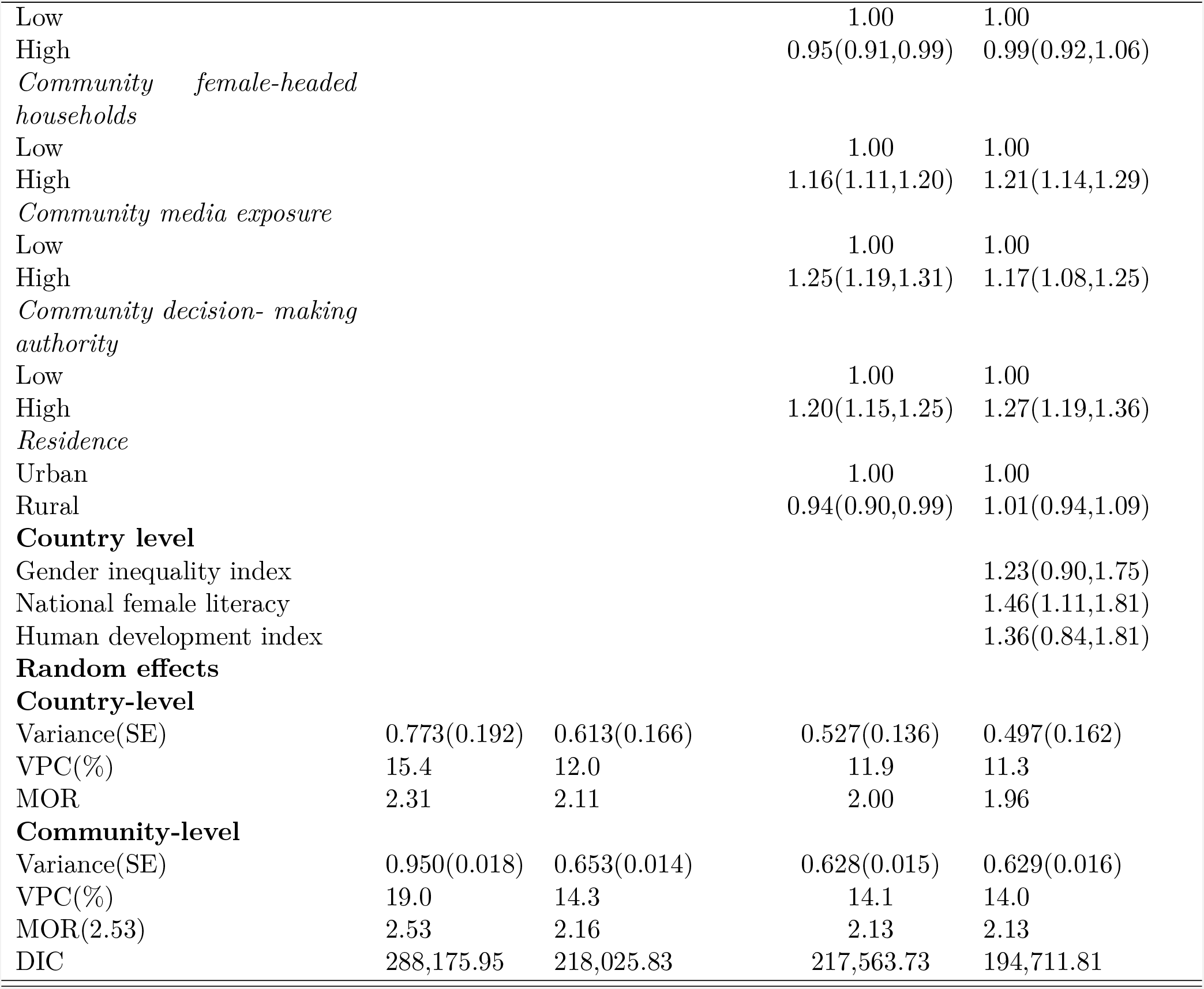
Gender relations and antenatal care analysis of pooled sample (N = 245, 955)

| Variable | Model 1 | Model 2 | Model 3 | Model 4 |
| --- | --- | --- | --- | --- |
| **Individual-level** |  |  |  |  |
| Intercept | 0.85 | 0.49 | 0.34 | 0.31 |
| *Maternal age* |  |  |  |  |
| <20 |  | 1.00 | 1.00 | 1.00 |
| 20-24 |  | 1.06(1.01,1.09) | 1.04(1.00,1.08) | 1.01(0.79,1.06) |
| 35-49 |  | 1.05(1.01,1.10) | 1.04(1.00,1.09) | 1.05(0.96,1.12) |
| *Educational status* |  |  |  |  |
| No education |  | 1.00 | 1.00 | 1.00 |
| Primary |  | 1.43(1.17,1.26) | 1.30(1.26,1.35) | 1.41(1.34,1.49) |
| Secondary/higher |  | 1.95(1.87,2.03) | 1.72(1.64,1.79) | 1.83(1.71,1.96) |
| *Household wealth* |  |  |  |  |
| First |  | 1.00 | 1.00 | 1.00 |
| Second |  | 1.22(1.17,1.26) | 1.16(1.12,1.20) | 1.12(1.06,1.19) |
| Middle |  | 1.49(1.43,1.55) | 1.33(1.28,1.39) | 1.40(1.31,1.49) |
| Fourth |  | 1.84(1.76,1.92) | 1.52(1.45,1.59) | 1.70(1.57,1.84) |
| Fifth |  | 2.82(2.67,2.96) | 2.06(1.94,2.18) | 2.41(2.18,2.65) |
| *Employment* |  |  |  |  |
| Unemployed |  | 1.00 | 1.00 | 1.00 |
| Self-employed(agric) |  | 0.99(0.94,1.02) | 1.02(0.98,1.06) | 1.03(0.97,1.10) |
| Formally employed |  | 1.20(1.17,1.24) | 1.18(1.14,1.21) | 1.24(1.18,1.29) |
| *Insurance coverage* |  |  |  |  |
| No |  | 1.00 | 1.00 | 1.00 |
| Yes |  | 1.30(1.21,1.38) | 1.27(1.19,1.36) | 1.63(1.47,1.82) |
| *Sexual autonomy* |  |  |  |  |
| No |  | 1.00 | 1.00 | 1.00 |
| Yes |  | 1.07(1.04,1.10) | 1.05(1.02,1.08) | 1.10(1.04,1.15) |
| *Negotiating sex* |  |  |  |  |
| No |  | 1.00 | 1.00 | 1.00 |
| Yes |  | 1.26(1.22,1.29) | 1.23(1.19,1.26) | 1.23(1.18,1.29) |
| *Condone violence* |  |  |  |  |
| No |  | 1.00 | 1.00 | 1.00 |
| Yes |  | 0.93(0.91,0.97) | 0.95(0.92,0.98) | 0.93(0.86,0.98) |
| **Community-level** |  |  |  |  |
| *Community education* |  |  |  |  |
| Low |  |  | 1.00 | 1.00 |
| Mediam |  |  | 1.49(1.41,1.57) | 1.75(1.58,1.93) |
| High |  |  | 1.60(1.49,1.72) | 1.21(1.61,2.01) |
| *Community distance problem* |  |  |  |  |
| Low |  |  | 1.00 | 1.00 |
| High |  |  | 0.91(0.87,0.95) | 0.84(0.78,0.90) |
| *Community money problem* |  |  |  |  |
| Low |  |  | 1.00 | 1.00 |
| High |  |  | 0.85(0.83,0.88) | 0.80(0.77,0.84) |
| *Community polygynous marriage* |  |  |  |  |
| Low |  |  | 1.00 | 1.00 |
| High |  |  | 0.95(0.91,0.99) | 0.99(0.92,1.06) |
| *Community female-headed households* |  |  |  |  |
| Low |  |  | 1.00 | 1.00 |
| High |  |  | 1.16(1.11,1.20) | 1.21(1.14,1.29) |
| *Community media exposure* |  |  |  |  |
| Low |  |  | 1.00 | 1.00 |
| High |  |  | 1.25(1.19,1.31) | 1.17(1.08,1.25) |
| *Community decision- making authority* |  |  |  |  |
| Low |  |  | 1.00 | 1.00 |
| High |  |  | 1.20(1.15,1.25) | 1.27(1.19,1.36) |
| *Residence* |  |  |  |  |
| Urban |  |  | 1.00 | 1.00 |
| Rural |  |  | 0.94(0.90,0.99) | 1.01(0.94,1.09) |
| **Country level** |  |  |  |  |
| Gender inequality index |  |  |  | 1.23(0.90,1.75) |
| National female literacy |  |  |  | 1.46(1.11,1.81) |
| Human development index |  |  |  | 1.36(0.84,1.81) |
| **Random effects** |  |  |  |  |
| **Country-level** |  |  |  |  |
| Variance(SE) | 0.773(0.192) | 0.613(0.166) | 0.527(0.136) | 0.497(0.162) |
| VPC(%) | 15.4 | 12.0 | 11.9 | 11.3 |
| MOR | 2.31 | 2.11 | 2.00 | 1.96 |
| **Community-level** |  |  |  |  |
| Variance(SE) | 0.950(0.018) | 0.653(0.014) | 0.628(0.015) | 0.629(0.016) |
| VPC(%) | 19.0 | 14.3 | 14.1 | 14.0 |
| MOR(2.53) | 2.53 | 2.16 | 2.13 | 2.13 |
| DIC | 288,175.95 | 218,025.83 | 217,563.73 | 194,711.81 |

**Table 4:**
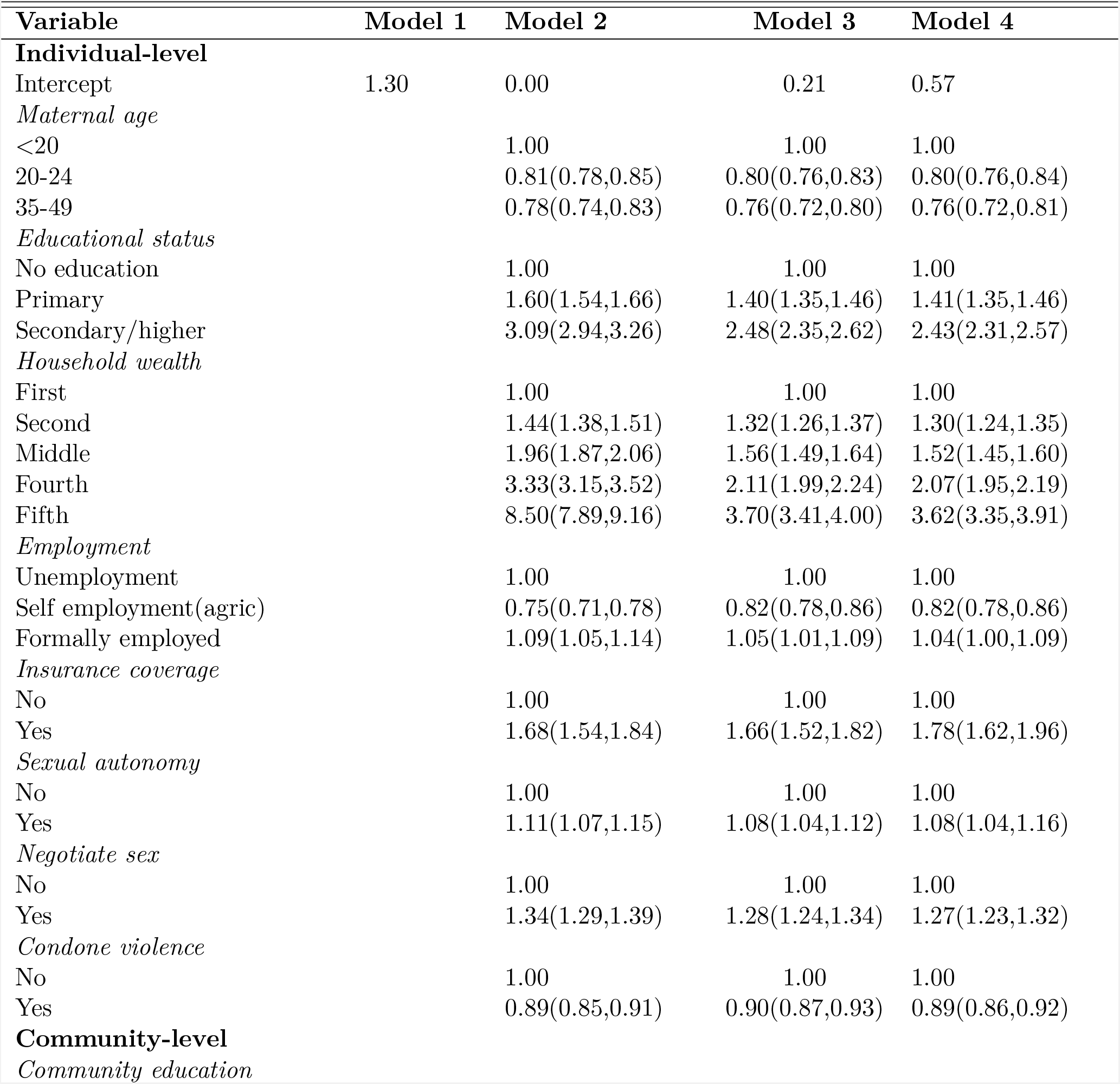
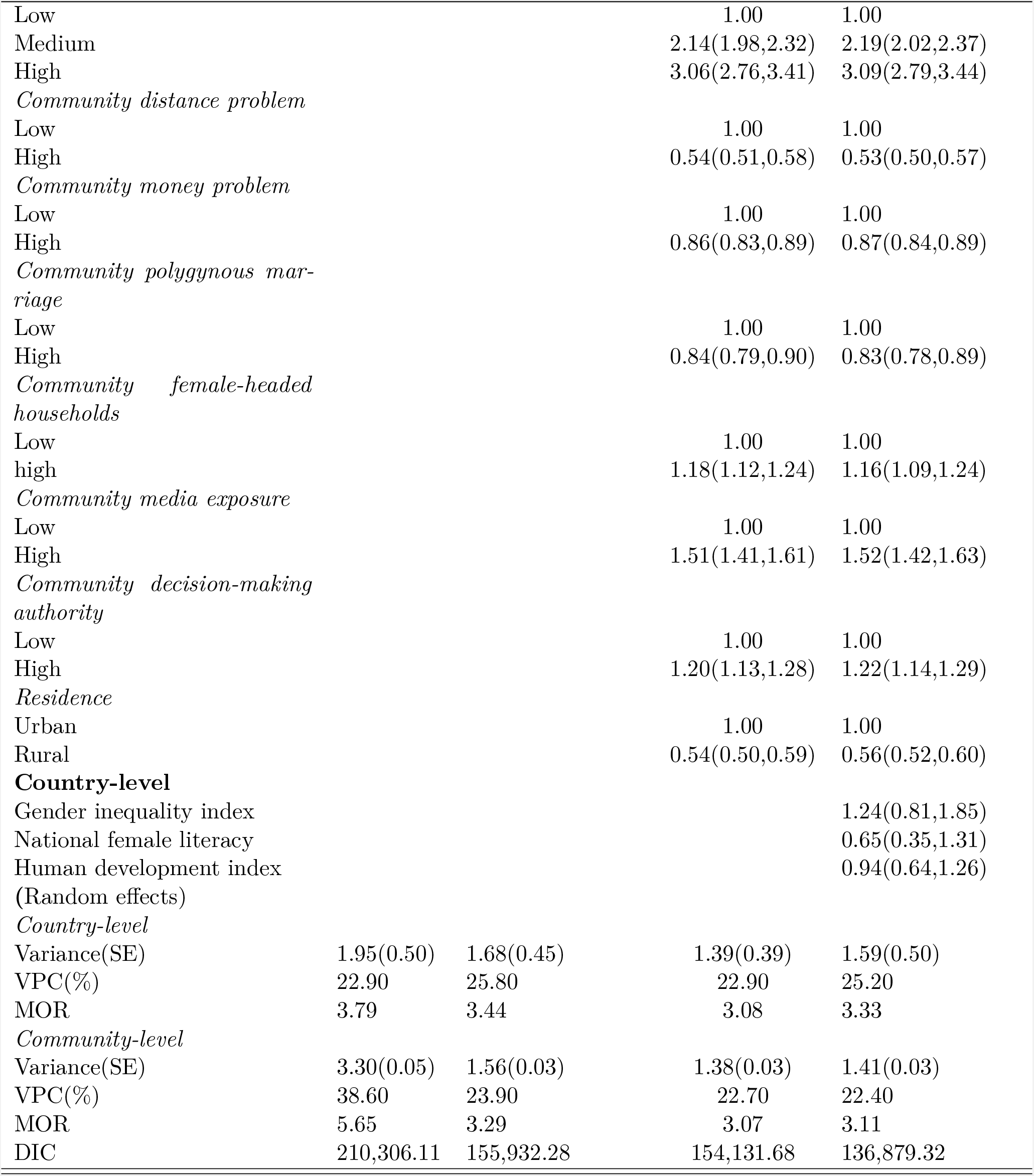
Gender relations and institutional delivery analysis pooled sample (N = 245, 955)

| Variable | Model 1 | Model 2 | Model 3 | Model 4 |
| --- | --- | --- | --- | --- |
| **Individual-level** |  |  |  |  |
| Intercept | 1.30 | 0.00 | 0.21 | 0.57 |
| *Maternal age* |  |  |  |  |
| <20 |  | 1.00 | 1.00 | 1.00 |
| 20-24 |  | 0.81(0.78,0.85) | 0.80(0.76,0.83) | 0.80(0.76,0.84) |
| 35-49 |  | 0.78(0.74,0.83) | 0.76(0.72,0.80) | 0.76(0.72,0.81) |
| *Educational status* |  |  |  |  |
| No education |  | 1.00 | 1.00 | 1.00 |
| Primary |  | 1.60(1.54,1.66) | 1.40(1.35,1.46) | 1.41(1.35,1.46) |
| Secondary/higher |  | 3.09(2.94,3.26) | 2.48(2.35,2.62) | 2.43(2.31,2.57) |
| *Household wealth* |  |  |  |  |
| First |  | 1.00 | 1.00 | 1.00 |
| Second |  | 1.44(1.38,1.51) | 1.32(1.26,1.37) | 1.30(1.24,1.35) |
| Middle |  | 1.96(1.87,2.06) | 1.56(1.49,1.64) | 1.52(1.45,1.60) |
| Fourth |  | 3.33(3.15,3.52) | 2.11(1.99,2.24) | 2.07(1.95,2.19) |
| Fifth |  | 8.50(7.89,9.16) | 3.70(3.41,4.00) | 3.62(3.35,3.91) |
| *Employment* |  |  |  |  |
| Unemployment |  | 1.00 | 1.00 | 1.00 |
| Self employment(agric) |  | 0.75(0.71,0.78) | 0.82(0.78,0.86) | 0.82(0.78,0.86) |
| Formally employed |  | 1.09(1.05,1.14) | 1.05(1.01,1.09) | 1.04(1.00,1.09) |
| *Insurance coverage* |  |  |  |  |
| No |  | 1.00 | 1.00 | 1.00 |
| Yes |  | 1.68(1.54,1.84) | 1.66(1.52,1.82) | 1.78(1.62,1.96) |
| *Sexual autonomy* |  |  |  |  |
| No |  | 1.00 | 1.00 | 1.00 |
| Yes |  | 1.11(1.07,1.15) | 1.08(1.04,1.12) | 1.08(1.04,1.16) |
| *Negotiate sex* |  |  |  |  |
| No |  | 1.00 | 1.00 | 1.00 |
| Yes |  | 1.34(1.29,1.39) | 1.28(1.24,1.34) | 1.27(1.23,1.32) |
| *Condone violence* |  |  |  |  |
| No |  | 1.00 | 1.00 | 1.00 |
| Yes |  | 0.89(0.85,0.91) | 0.90(0.87,0.93) | 0.89(0.86,0.92) |
| **Community-level** |  |  |  |  |
| *Community education* |  |  |  |  |
| Low |  |  | 1.00 | 1.00 |
| Medium |  |  | 2.14(1.98,2.32) | 2.19(2.02,2.37) |
| High |  |  | 3.06(2.76,3.41) | 3.09(2.79,3.44) |
| *Community distance problem* |  |  |  |  |
| Low |  |  | 1.00 | 1.00 |
| High |  |  | 0.54(0.51,0.58) | 0.53(0.50,0.57) |
| *Community money problem* |  |  |  |  |
| Low |  |  | 1.00 | 1.00 |
| High |  |  | 0.86(0.83,0.89) | 0.87(0.84,0.89) |
| *Community polygynous marriage* |  |  |  |  |
| Low |  |  | 1.00 | 1.00 |
| High |  |  | 0.84(0.79,0.90) | 0.83(0.78,0.89) |
| *Community female-headed households* |  |  |  |  |
| Low |  |  | 1.00 | 1.00 |
| high |  |  | 1.18(1.12,1.24) | 1.16(1.09,1.24) |
| *Community media exposure* |  |  |  |  |
| Low |  |  | 1.00 | 1.00 |
| High |  |  | 1.51(1.41,1.61) | 1.52(1.42,1.63) |
| *Community decision-making authority* |  |  |  |  |
| Low |  |  | 1.00 | 1.00 |
| High |  |  | 1.20(1.13,1.28) | 1.22(1.14,1.29) |
| *Residence* |  |  |  |  |
| Urban |  |  | 1.00 | 1.00 |
| Rural |  |  | 0.54(0.50,0.59) | 0.56(0.52,0.60) |
| **Country-level** |  |  |  |  |
| Gender inequality index |  |  |  | 1.24(0.81,1.85) |
| National female literacy |  |  |  | 0.65(0.35,1.31) |
| Human development index |  |  |  | 0.94(0.64,1.26) |
| (Random effects) |  |  |  |  |
| *Country-level* |  |  |  |  |
| Variance(SE) | 1.95(0.50) | 1.68(0.45) | 1.39(0.39) | 1.59(0.50) |
| VPC(%) | 22.90 | 25.80 | 22.90 | 25.20 |
| MOR | 3.79 | 3.44 | 3.08 | 3.33 |
| *Community-level* |  |  |  |  |
| Variance(SE) | 3.30(0.05) | 1.56(0.03) | 1.38(0.03) | 1.41(0.03) |
| VPC(%) | 38.60 | 23.90 | 22.70 | 22.40 |
| MOR | 5.65 | 3.29 | 3.07 | 3.11 |
| DIC | 210,306.11 | 155,932.28 | 154,131.68 | 136,879.32 |

I report the posterior odds ratios and 95% Bayesian credible intervals (CrI) for each of the variables in all models except for the null. Statistical significance is determined by non-inclusion of “1” in the 95% (CrI). Both the fixed and random effects are reported. Fixed effects are the average associations of individual, community and country-level variables on maternal health care and these are represented as odds ratios and 95% credible intervals. Random effects are measures of variations in maternal health care use across communities and countries. To measure random effects, I used the variance partition coefficient (VPC) which measures the proportion of variation in the outcome variable that occurs between groups versus the total variation present. Higher VPC values show that a greater share of total variation in the out come variables is attributable to higher level membership (Finch et al., 2019).

### Gender relations, women empowerment, and antenatal care

Table 3 presents the results of 4 models analyzing the odds of having adequate antenatal care visits during pregnancy. Model 2 which represents individual-level variables, indicates that maternal age, educational status,employment status, sexual autonomy, insurance coverage and condoning violence are all associated with having adequate antenatal care visits. Women aged 20 years and above, those who are educated up to primary level or higher, who are in employment, are able to refuse or negotiate sex and have health coverage are more likely to have of 4 or more antenatal care visits to health facilities during pregnancy. It has also been established that women who condone violence are significantly less likely to have adequate antenatal care. Model 3 includes contextual factors at the community level. Community education, community media exposure and community decision-making authority are associated with 4 or more antenatal care visits. This means that women living in communities where a higher proportion of women have an education and those living in communities where a higher proportion have media exposure are more likely to use antenatal care services. The same is true for communities with a high proportion of women having decision-making authorities within their families.

As expected, distance and money-related barriers to access health facilities are negatively associated with ANC visits. Women residing in clusters in which a high proportion of women consider the lack of money and distance to be major problems in accessing healthcare facilities were less likely to use antenatal care services. Inequalities in gender relations variables including the proportion of female-headed households and polygynous marriages in a community show interesting results. Women who live in communities with high proportions of polygynous marriages are less likely to have adequate antenatal care while those who live in communities with high proportions of female-headed households are more likely to have adequate antenatal care use. All the individual level variables that were associated with antenatal care use in the second model remained so even after controlling for the effects of community-level variables.

Model 4 includes contextual factors at both country and community levels. It shows that only maternal age at the individual level, community polygynous marriages and residence at the community level lose significance when country-level variables are factored in. However, apart from the female literacy rate, the other country-level variables are not significantly associated with antenatal care use. For female literacy, the results show that a one unit increase in female employment, increases the odds of antenatal care use by a factor of 1.46.

Table 3 also reports the variance partition coefficients (VPCs), which show that the proportion of unexplained variation in antenatal care attributable to the community and country level factors is significantly large, 19.0% and 15.4% respectively. Antenatal care variations across communities and countries remained significant even in model 4 which controls for all the factors. The median odds ration (MOR) for the unadjusted model at both community and country-levels are substantial, 2.52 and 2.31 respectively. These values confirm the importance contextual factors in explaining the odds of having adequate antenatal care visits.

Model comparison denoted by the values of the DIC shows an improvement in model fit as the variables at different levels are being added. This means a complete three-level model is a considerably better predictor of adequate antenatal care compared to other models.

### Gender relations, women empowerment, and institutional delivery

Table 4 presents analyses of the odds of having institutional delivery among women in sub-Saharan Africa. The results show similar patterns as the antenatal care uptake. All individual level variables are consistently associated with institutional delivery even after controlling for community and country-level variables. However, it is interesting to note that for maternal age, women who are more than 20 years are less likely to deliver in health facilities when the opposite was the case for antenatal care visits. Additionally, women who are employed in agriculture have consistently reduced odds of delivering in institutional facilities compared to those who are unemployed. All community-level variables are associated with institutional delivery including community polygamous marriages and place of residence which unlike in the antenatal care model, they are consistently negatively associated with institutional delivery. The country-level variables are all not significantly associated with facility delivery.

The VPC indicates that unlike antenatal care, the variations in the odds of institutional delivery are explained by the contextual factors more than individual factors. This is the case even after controlling for all the factors. The MOR from the variances are also very high for both the country level (MOR = 3.79) and the community level (MOR = 5.65) indicating greater contextual-level variations in institutional delivery.

The DIC which which reports model comparison shows an improvement in model fit as the variables are different levels are being added. This means a complete three-level model is a considerably better predictor of institutional delivery compared to other models.

## Discussion

This study sought to examine the influence of gender relations and women empowerment on maternal health care utilization and the role played by contextual factors in this relationship. Gender relations is are social phenomena that are influenced by social, cultural and economic factors (Sado et al., 2014) and are in turn reflected mainly by women’s interactions with intimate partners, households, community and society at large (Kane et al., 2016). Maternal health care in sub-Saharan Africa is akin to sexual and reproductive health which as Organization et al. (2007) describes, is shaped by the nature of intimate relationships, family structures, community institutions and relations. To adequately explain variations in maternal healthcare utilization, it is important to capture this complexity in the analysis. I used three-level multilevel logistic regression models to represent individual, community and country level measures of gender relations and other inter-sectional categories. The findings of this study provides evidence of direct linkages between these measures and the two most basic indicators of maternal health care uptake, antenatal care and institutional delivery.

The study deepens our understanding of the importance of contextual gendererd factors in maternal healthcare utilization. In particular, the study found that women who live in communities where there are high proportions of women with female-headed households, decision making authority, low polygamy prevalence are more likely to utilize maternal healthcare in sub-Saharan Africa. Other community level factors that are significantly associated with adequate utilization of maternal healthcare include education, media exposure, distance to health facilities and money problems.

The study finds evidence of substantial clustering at both the community and country levels. Both the VPC and MOR confirm that contextual level factors are responsible for more variations in institutional delivery care than individual-level variables. Individual-level factors still explains greater variation in antenatal care coverage albeit with significant contextual-level variability. Clustering evidence indicates the presence of homogeneity among women from the same communities and countries, suggesting that they are shaped by common sociocultural factors.

Gender relations factors, female-headed household, decision-making authority and polygamous marriages were also measured at the individual-level and were found to be significantly associated with maternal healthcare. I also included sexual autonomy, ability to negotiate sex and condoning violence at the individual level and these were found to be significantly associated with maternal healthcare utilization. Control variations, including education, employment, media exposure, autonomy and being from a are predictors of consistent use of maternal health care in sub-Saharan Africa. There are many studies that have reported significant relationships between several gender-related factors and use of maternal health care in sub-Saharan Africa, some of which corroborate our results albeit in different contexts (Adjiwanou and LeGrand, 2014; Gage, 2007; Onah et al., 2006; Ononokpono et al., 2013; Stephenson et al., 2006). However, these studies either use individual countries, a few selected countries or do not focus on broader contextual influences of gender-related factors on maternal health care.

The results are also corroborated by (Ahmed et al., 2010) who use multiple countries and found empowerment, education and empowerment to be significantly associated with maternal health care, measured as contraceptive use, skilled birth attendance and attending at least four antenatal care visits in developing countries. Adjiwanou et al. (2018) also finds a significant relationship between education and maternal health care utilization.

Maternal health, throughout the continuum of pregnancy care, including attending a recommended number of antenatal visits, having skilled delivery care and postnatal checkups is central to woman’s health which leads to reduced maternal mortality and have positive implications for society as a whole. The results raise important implications across the SSA society as to the social mechanisms that contribute to the observed associations. It shows that health care disparities are embedded within the social and cultural fabric and is based on gender relations and socio-economic status. The pathways between high socio-economic status and healthcare utilization are obvious as the person who has wealth and is educated is most likely to reside in places which are proximal to health facilities and is also most likely to have health insurance or afford out-of-pocket payments for healthcare (Baum et al., 1999; Cutler et al., 2008; Leive and Xu, 2008; Myburgh et al., 2005). Gender relations in the form of lack of decision-making authority among women, affects levels of emotional, sexual and physical well-being as well as mobility to healthcare facilities and hence poor health outcome (Krug et al., 2002; Matthews et al., 1999; Steele et al., 2001; Steele and Goldstein, 2006).

National female literacy at the community and country levels that is found to be significantly associated with maternal health care buttresses notions of the impact of up-stream factors in individual level decision-making processes. This would be the result of structural factors embedded within local communities and broader social institutions existing at the country and international level.In fact, individual level determinants of maternal healthcare utilization may just be symptoms of structural factors within local communities and broader social institutions. For example, the influence of decision-making authority on maternal healthcare operationalized at an individual level have often been discussed within the framework of dominant and broader masculinity ideologies or cultural beliefs in particular spaces (Say and Raine, 2007). The same may be true with health systems whose dysfunctionality may be a direct consequence of political and governance systems far removed from their level of operation. Indeed some studies have found evidence of the significant role played by structural factors in influencing maternal healthcare utilization in sub-Saharan Africa.

Limitations of the study include the use of pooled analysis which combined the 35 countries of sun-Saharan Africa. It is probable that effect sizes resulting from the pooled analysis may not represent what pertains within individual countries included. Also, the constitution of community level variables which was done by aggregating individual level variables using the PSUs may potentially have created atomistic fallacy (Diez, 2002) whereby inferences at the high level are made using lower level data. Recall bias is also one of the potential problems although it was minimized by the focus on exclusively the most recent birth of the five years prior to the survey.

The strengths of this study lies in the use of several factors to measure gender relations at three-levels, providing a cross-national evidence of the magnitude of their associations with use maternal health care. It is also worth noting that unlike some previous studies (Ononokpono et al., 2013) that have relied on one variable to represent maternal healthcare utilization, this study uses the whole continuum beginning with antenatal care up to postnatal check-ups for mothers and newborn children. The use of Bayesian MCMC estimation which minimizes bias to estimation especially in multilevel models with countries occupying the highest level in the analysis (Bryan and Jenkins, 2016; Stegmueller, 2013). This provides a better separations of the effects of individual and contextual factors on women’s utilization of MHC, which is critical for the implementation of policy strategies aimed at bolstering the use of MHC services especially in low resource countries. Isolating the effects of contextual conditions provides a better platform for theoretical developments that are crucial for understanding the relationships between the broader social structure and health outcomes. This may ultimately help us explain why there are protracted disparities in health and healthcare outcomes in sub-Saharan Africa.

## Conclusion

This paper addressed the impact of gender relations on maternal healthcare utilization in sub-Saharan Africa. I used several variables at the individual, community and country-level to represent gender relations. The results show that in addition to several individual level factors, female-headed households and community decision-making authority are significant predictors of both antenatal care and institutional delivery care. Community education, community media exposure and community distance to health facilities are found to be significant predictors of maternal healthcare utilization.

The findings from this study also generally attributes cross-national variations in maternal healthcare to contextual factors. The study found that apart from specific individual, community and country levels factors that are found to be associated with indicators of maternal healthcare, the findings indicate that contextual factors are most important in explaining variations in maternal healthcare. These findings help to emphasize the importance of contextual factors in terms of understanding as well as bolstering maternal healthcare utilization in sub-Saharan Africa.

## Data Availability

All data produced are available online at: //dhsprogram.com/data/available-datasets.cfm

